# Accuracy and Precision of 3D Optical Imaging for Body Composition and their Associations to Metabolic Markers by Age, BMI, and Ethnicity

**DOI:** 10.1101/2022.11.02.22281819

**Authors:** Michael C. Wong, Jonathan P. Bennett, Brandon Quon, Lambert T. Leong, Isaac Y. Tian, Yong E. Liu, Nisa N. Kelly, Cassidy McCarthy, Dominic Chow, Sergi Pujades, Andrea K. Garber, Gertraud Maskarinec, Steven B. Heymsfield, John A. Shepherd

## Abstract

**Background:** Recent three-dimensional optical (3DO) imaging advancements have provided a more accessible, affordable, and self-operating opportunities for assessing body composition. 3DO is accurate and precise compared to clinical measures measured by dual-energy X-ray absorptiometry (DXA) in diverse study samples. However, the accuracy and precision of an overall 3DO body composition model in specific subgroups is unknown. Therefore, this study aimed to evaluate 3DO’s accuracy and precision by subgroups of age, body mass index (BMI), and ethnicity/race.

**Methods:** A cross-sectional analysis was performed using the Shape Up! Adults study. Each participant received duplicate 3DO and DXA scans. 3DO meshes were digitally registered and reposed using Meshcapade to standardize the vertices and pose. Principal component analysis was performed on the registered 3DO meshes to orthogonalize and reduce the dimensionality of the data. The resulting principal components estimated DXA whole-body and regional body composition using stepwise forward linear regression with five-fold cross-validation. Duplicate 3DO and DXA scans were used for test-retest precision. Student’s t-test was performed between 3DO and DXA by subgroup to determine significant differences. One-way ANOVA determined if intra-group precision had significant differences. P-value < 0.05 was considered statistically significant.

**Results:** Six hundred thirty-four participants (females = 346) had completed the study at the time of the analysis. 3DO total fat mass (FM) and fat-free mass (FFM) in the entire sample achieved R^2^s of 0.94 and 0.92 with RMSEs of 2.91 kg and 2.76 kg, respectively, in females and similarly in males. 3DO total FM and FFM achieved a %CV (RMSE) of 1.76% (0.44 kg) and 1.97% (0.44 kg), while DXA had a %CV (RMSE) of 0.98% (0.24 kg) and 0.59% (0.27 kg), respectively, in females and similarly in males. There were no mean differences by age group (p-value > 0.068). However, there were mean differences for underweight females, NHOPI females and males, and Asian and black females (p-value < 0.038). There were no significant differences among the subgroups for precision (p-value > 0.109).

**Conclusion:** A single 3DO body composition model derived from a highly-stratified dataset performed well against DXA with minimal differences detected for accuracy and precision. Adjustments to specific subgroups may be warranted to improve the accuracy in those that had significant differences. Nevertheless, 3DO produced accurate and precise body composition estimates that can be used on diverse populations.

## INTRODUCTION

Obesity is defined by having excess adiposity and has been a growing epidemic for the past few generations (1-3). This excess adiposity is linked to the development of cardiovascular disease, type-II diabetes, and up to 20% of cancers (4-6). Body mass index (BMI) is generally used to classify obesity. However, BMI only uses height and weight and does not account for muscle or fat mass, and therefore, a poor method for nutritional assessment on an individual level. Instead, body composition methods have been developed to quantify fat mass (FM) and fat-free mass (FFM) in order to provide a more thorough health assessment (7).

Magnetic resonance and computer tomography (CT) are among criterion methods for body composition (8, 9). However, these methods are expensive, require trained/certified technicians, use large radiation doses (CT), lack accessibility to people outside clinical settings, and are not recommended for routine use. Dual-energy X-ray absorptiometry (DXA), air displacement plethysmography (ADP), and bioelectric impedance analysis (BIA) are among the more accessible modalities developed for body composition, but even they have limitations such as requiring trained technicians (DXA and ADP), using ionizing radiation (DXA), many built-in physiological assumptions (ADP and BIA), inconsistent accuracy and precision by make and model (BIA), and not providing regional/compartmental compositions (ADP) (10-12). The ideal method would include total and regional composition, free of ionizing radiation, self-operating, and accessible to many.

In the last two decades or so, three-dimensional optical (3DO) imaging has been deeply explored as a health assessment method. 3DO scanners generally output a detailed 3D mesh that represents the person’s body shape and automated anthropometric estimates (i.e., circumferences, lengths, surface areas, and volumes) (13, 14). During the early days, researchers validated the accuracy and precision of the automated anthropometry to criterion methods (e.g., tape measurements to circumferences/lengths, underwater weighing or ADP to total volume) (15, 16). 3DO waist circumference, hip circumference, and waist-hip-ratio were highly associated to metabolic markers and improved predictions for metabolic syndrome (17, 18). Researchers also showed that these automated anthropometric estimates, where some would be difficult and tedious to obtain manually, were predictive of DXA FM and FFM (19-22).

Since 3DO anthropometry was as good, if not better than manual measurements, as health descriptors, researchers attempted to created better shape descriptors using the 3DO mesh to utilize the entire body. They showed that body shape descriptors by the 3DO mesh were more predictive of body composition as compared to DXA and were correlated to metabolic biomarkers (23). Since then, the methodology to obtain these body shape descriptors have improved by incorporating automated processing methods, agnostic models across multiple 3DO scanners, pose-independent models, and a 2D to 3D pipeline (24-27). Although much has been accomplished in this field in a short amount of time, 3DO body composition has not been evaluated at the subgroup level. In order for 3DO to be accepted as a viable body composition method, 3DO must be scrutinized at a more minute scale. The hypothesis was that 3DO body composition does not differ from DXA by subgroups of age, BMI, or ethnicity/race. The objective of this study was to evaluate the accuracy and precision by subgroups of age, BMI, and ethnicity/race as compared to DXA.

## METHODS

### Study Design

Shape Up! Adults was a cross-sectional study of healthy adults (NIH R01 DK109008, ClinicalTrials.gov ID NCT03637855). This study was designed to investigate the associations between body shape and composition with various health markers. Participants underwent whole-body 3DO scans, dual energy X-ray absorptiometry (DXA) scans, blood serum tests, and functional tests for hand grip and knee extension strength.

### Participants

Participants were recruited at Pennington Biomedical Research Center (PBRC), University of Hawaii Cancer Center (UHCC), and University of California, San Francisco (UCSF). All participants provided informed consent. The study protocol was approved by the Institutional Review Boards (IRBs) at PBRC (IRB study #2016-053), UCSF (IRB #15-18066), and the University of Hawaii Office of Research Compliance (UH ORC, CHS #2017-01018). Volunteers were pre-screened over the phone and were deemed ineligible if they were pregnant, breastfeeding, had missing limbs, non-removable metal, previous body-altering surgery (e.g., breast augmentation, liposuction), hair that could not be contained in a swim cap, were unable to stand still for one minute, or unable to lay still for three minutes. Pretesting preparations included an eight-hour fast (water and prescribed medications were allowed) and no strenuous exercise 24 hours prior to the study visit. Participants were stratified by age (18-39, 40-59, ≥60 years), ethnicity (non-Hispanic white (white), non-Hispanic black (black), Hispanic, Asian, and Native Hawaiian or Pacific Islander (NHOPI)), sex, and BMI (<18.5, 18.5-25, 25-30, >30 kg/m2). Height and weight were measured on a SECA 274 Stadiometer (SECA GmbH, Hamburg, Germany).

### Metabolic Blood Biomarkers

Each participant had 40 ml of blood drawn intravenously by a certified, trained phlebotomist. Prior to blood draw, participants fasted for at least eight hours with the exception of water and prescription medication. Blood samples were placed on ice and processed within four hours into plasma, whole blood, buffy coat components, and serum. Samples were stored in -80°C at each study site until sent to PBRC for batched analysis. Serum chemistry panels were assayed by a DXC600 instrument (Beckman Coulter, Inc.; Brea, CA). LDL cholesterol was calculated as [total cholesterol] – [HDL cholesterol] – [triglycerides]/5 (all values in mg/dL) as described by Friedewald, et al. (28). Insulin was measured by immunoassay on an Immulite 2000 platform (Siemens Corporation; Washington, DC). Present analysis used fasting glucose, HbA1c, total cholesterol, LDL and HDL cholesterol, serum triglycerides, and insulin measurements.

### Strength Measurements

Isokinetic leg strength was measured using a Biodex System 4 (Biodex Medical Systems Inc., Shirley, NY, USA) or HUMAC NORM (Computer Sports Medicine Inc., Stoughton, MA, USA) dynamometer. Only the dominant leg was used for testing. The dominant leg was defined as the leg one would use to kick a ball. Participants were fastened into the seat with a seat belt and a Velcro strap over their dominant leg to isolate movement. The dynamometer was adjusted accordingly to accommodate the participant’s upper and lower leg length. The participants then practiced at an endurance of 50% of maximal effort for isokinetic testing until they were comfortable with the procedures. For the isokinetic measurement, resistance was set at 60° per second. After the practice round, participants performed a set of five repetitions at maximal effort. Peak torque was recorded as the maximum torque (in units of Newton-meters) achieved during the repetitions.

### Dual-Energy X-Ray Absorptiometry

Participants received duplicate whole-body DXA scans with a Hologic Discovery/A or Horizon system (Hologic Inc., Marlborough, Massachusetts) according to International Society for Clinical Densitometry guidelines (29) with repositioning. Participants were scanned once, asked to get off the table, and laid back on the table for the second scan. A phantom was scanned daily for quality control and used for calibration across sites. All scans were analyzed at UHCC by a single certified technologist using Hologic Apex version 5.6 with the National Health and Nutrition Examination Survey Body Composition Analysis calibration option disabled (30).

### 3D Optical Surface Scans

Participants changed into form-fitting tights, a swim cap, and a sports bra if female. Duplicate 3DO surface scans were taken on the Fit3D ProScanner version 4.x (Fit3D Inc., San Mateo, California) within ten minutes of each other and with repositioning. Participants grasped telescoping handles on the scanner platform and stood upright with shoulders relaxed and arms positioned straight and abducted from their torso. The platform rotates once around and takes approximately 45 seconds for the completion of the scan. Final point clouds were converted to a mesh connected by triangles with approximately 300,000 vertices and 600,000 faces to represent the body shape (20).

3DO meshes were sent to Meshcapade (Meshcapade GmbH, Tubingen, Germany) for registration and to be digitally reposed. Their algorithm registers each mesh to a 110,000-vertex template with full anatomical correspondence. This means each vertex corresponds to a specific anatomical location across all registered meshes. All meshes were digitally reposed to a T-pose, where the person was standing straight, arms were brought horizontal and in plane with the body, and arms and legs were straightened (26, 31).

### Statistical Analysis

Since vertices in the 3D meshes are highly correlated to neighboring vertices, principal component analysis (PCA) was performed on the registered meshes to orthogonalize and reduce the dimensionality of the data so that fewer variables are needed to describe the data’s variance (23). The resulting outputs were principal components (PCs) that could be used for analysis.

Stepwise forward linear regression with five-fold cross-validation was used to predict DXA body composition with the PCs. The independent variables were the PCs and demographics (i.e., age and BMI), while the dependent variables were DXA whole-body and regional body composition measures. PC-only models were created first and then adjusted with age and BMI as potential covariates for the PC + demographics models. Results were reported with the coefficient of determination (R^2^) and root mean square error (RMSE).

Test-retest precision, also known as short-term precision, was performed on the duplicate DXA and 3DO meshes. The percent coefficient of variation (%CV) and RMSE were used to quantify the test-retest precision defined by Gluer et al. (32). Precision between subgroup was calculated similarly and compared within the strata of age, BMI, and ethnicity/race with a one-way ANOVA. A p-value < 0.05 was considered statistically significant.

The accuracy of 3DO total FM, FFM, and %fat was evaluated at the subgroup level with the stratifications mentioned previously. Mean differences were calculated between 3DO PC + demographic estimates and DXA (3DO – DXA). Paired Student’s t-test determined if the differences were statistically significant (p-value < 0.05). If significant, the percent mean difference would be calculated [(DXA – 3DO) / DXA * 100]. Percent mean differences under 2% was considered small, 2-5% moderate, and > 5% large.

Pearson correlations were performed between total FM, FFM, and %fat by 3DO and DXA to metabolic blood markers and leg strength. A bootstrap confidence interval was implemented to determine if the strength of the association between 3DO and DXA were significantly different (33). P-values < 5.6E-5 were considered significant correlations after Bonferroni adjustment (Bonferroni, 0.05/900) (34). The metabolic marker and strength sub-analysis was performed in SAS version 9.4 for Windows (SAS Institute, Cary, North Carolina). All other analyses were performed in R version 4.2.1 (R Core Teams).

## RESULTS

Six hundred thirty-four participants had completed the study at the time of the analysis (Table1). Average age of females and males was 46.6 ± 15.9 and 43.5 ± 15.8 years, respectively. Average BMI of females and males was 27.1 ± 7.35 and 28.2 ± 5.94 kg/m^2^, respectively. Ethnicity/racial breakdown for females was 23.7% Asian, 20.8% non-Hispanic Black, 13.9% Hispanic, 8.4% NHOPI, and 33.2% non-Hispanic black. Ethnicity/racial breakdown for males was 25.3% Asian, 22.2% non-Hispanic Black, 11.5% Hispanic, 5.9% NHOPI, and 35.1% non-Hispanic black. Average DXA %fat was 34.1% ± 7.57% for females and 22.9% ± 6.6% for males.

**Table 1.** Overall sample characteristics

|  | **Females**<br>**(N=346)** | **Males**<br>**(N=288)** |
| --- | --- | --- |
| **Age (years)** |  |  |
| Mean (SD) | 46.6 (15.9) | 43.5 (15.8) |
| Median [Min, Max] | 47.0 [18.0, 89.0] | 42.0 [18.0, 79.0] |
| **Ethnicity** |  |  |
| Asian | 82 (23.7%) | 73 (25.3%) |
| Black | 72 (20.8%) | 64 (22.2%) |
| Hispanic | 48 (13.9%) | 33 (11.5%) |
| NHOPI | 29 (8.4%) | 17 (5.9%) |
| White | 115 (33.2%) | 101 (35.1%) |
| **Height (cm)** |  |  |
| Mean (SD) | 162 (6.88) | 176 (7.61) |
| Median [Min, Max] | 162 [144, 190] | 176 [147, 202] |
| **Weight (kg)** |  |  |
| Mean (SD) | 71.5 (20.3) | 87.4 (20.5) |
| Median [Min, Max] | 67.6 [35.4, 153] | 84.1 [40.6, 174] |
| **BMI (kg/m<sup>2</sup>)** |  |  |
| Mean (SD) | 27.1 (7.35) | 28.2 (5.94) |
| Median [Min, Max] | 26.4 [14.2, 53.1] | 27.8 [17.0, 52.6] |
| **Total Fat Mass (kg)** |  |  |
| Mean (SD) | 25.4 (11.9) | 20.9 (10.0) |
| Median [Min, Max] | 24.1 [6.30, 72.7] | 19.5 [5.01, 66.5] |
| **Total FFM Mass (kg)** |  |  |
| Mean (SD) | 46.1 (9.54) | 66.6 (12.4) |
| Median [Min, Max] | 44.2 [28.6, 80.4] | 65.5 [33.9, 108] |
| **Percent Fat (%)** |  |  |
| Mean (SD) | 34.1 (7.57) | 22.9 (6.60) |
| Median [Min, Max] | 34.4 [12.6, 53.3] | 23.1 [9.03, 38.6] |
| **VAT (kg)** |  |  |
| Mean (SD) | 0.45 (0.30) | 0.49 (0.28) |
| Median [Min, Max] | 0.37 [0.05, 1.37] | 0.42 [0.16, 1.64] |
| **Glucose (mg/dL)** |  |  |
| Mean (SD) | 93.0 (18.2) | 94.8 (18.8) |
| Median [Min, Max] | 89.0 [65.0, 230] | 92.0 [64.0, 279] |
| **Insulin (mg/dL)** |  |  |
| Mean (SD) | 10.7 (7.41) | 14.9 (33.6) |
| Median [Min, Max] | 9.05 [2.10, 45.3] | 10.3 [2.00, 515] |
| **HOMA-IR (mg/dL)** |  |  |
| Mean (SD) | 2.57 (2.09) | 3.85 (11.3) |
| Median [Min, Max] | 2.00 [0.39, 16.6] | 2.43 [0.41, 174] |
| **HbA1c (%)** |  |  |
| Mean (SD) | 0.53 (0.12) | 0.59 (0.15) |
| Median [Min, Max] | 0.51 [0.28, 1.62] | 0.55 [0.36, 1.88] |
| **Triglycerides (mg/dL)** |  |  |
| Mean (SD) | 92.9 (57.5) | 106 (78.3) |
| Median [Min, Max] | 76.0 [23.0, 417] | 81.0 [29.0, 538] |
| **LDL Cholesterol (mg/dL)** |  |  |
| Mean (SD) | 115 (31.0) | 112 (32.3) |
| Median [Min, Max] | 113 [37.1, 210] | 110 [44.2, 212] |
| **HDL Cholesterol (mg/dL)** |  |  |
| Mean (SD) | 63.5 (17.0) | 52.9 (13.8) |
| Median [Min, Max] | 60.7 [32.2, 141] | 51.9 [17.6, 99.1] |
| **Isokinetic Knee Extension (N m)** |  |  |
| Mean (SD) | 87.0 (33.4) | 153 (59.6) |
| Median [Min, Max] | 82.5 [18.8, 223] | 147 [22.2, 339] |

Abbreviations: BMI (body mass index), HbA1c (glycated hemoglobin), HDL (high-density lipoprotein), HOMA-IR (Homeostatic Model Assessment for Insulin Resistance), LDL (low-density lipoprotein), NHOPI (Native Hawaiian and other Pacific Islanders), VAT (visceral adipose tissue)
Body composition values measured from DXA.

Sex-specific statistical shape models were created with the registered and reposed 3DO meshes. Three PCs captured 95% of the shape variance in each of the female and male shape models. 3DO PC-only equations were highly correlated to DXA body composition values (Table 2). 3DO total FM and FFM in females achieved R^2^s of 0.94 and 0.92 with RMSEs of 2.91 kg and 2.76 kg, while males achieved R^2^s of 0.94 and 0.94 with RMSEs of 3.04 kg and 2.97 kg, respectively. Female and male 3DO %fat had moderate correlations to DXA (R^2^; 0.75 and 0.73, RMSE; 3.82% and 3.31%, respectively). Regional 3DO FM and FFM estimates (i.e., arms, legs, and trunk) had strong correlations to DXA (R^2^ range: 0.79 – 0.95, RMSE range: 0.27 – 1.86 kg) for females and males. After possible adjustments for age and BMI as covariates (PC + demographics), there were marginal improvements to the R^2^s and RMSEs.

**Table 2.**
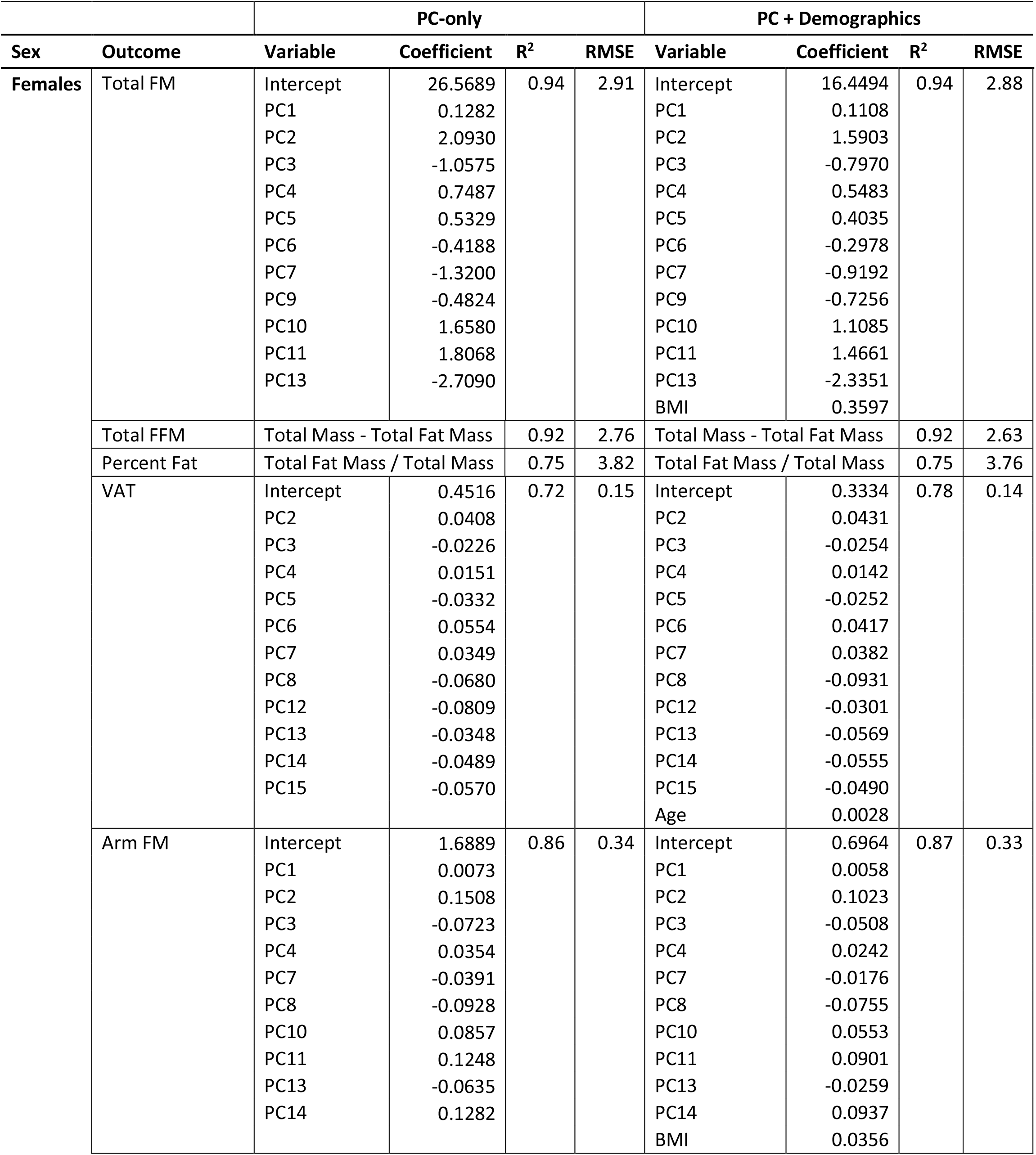
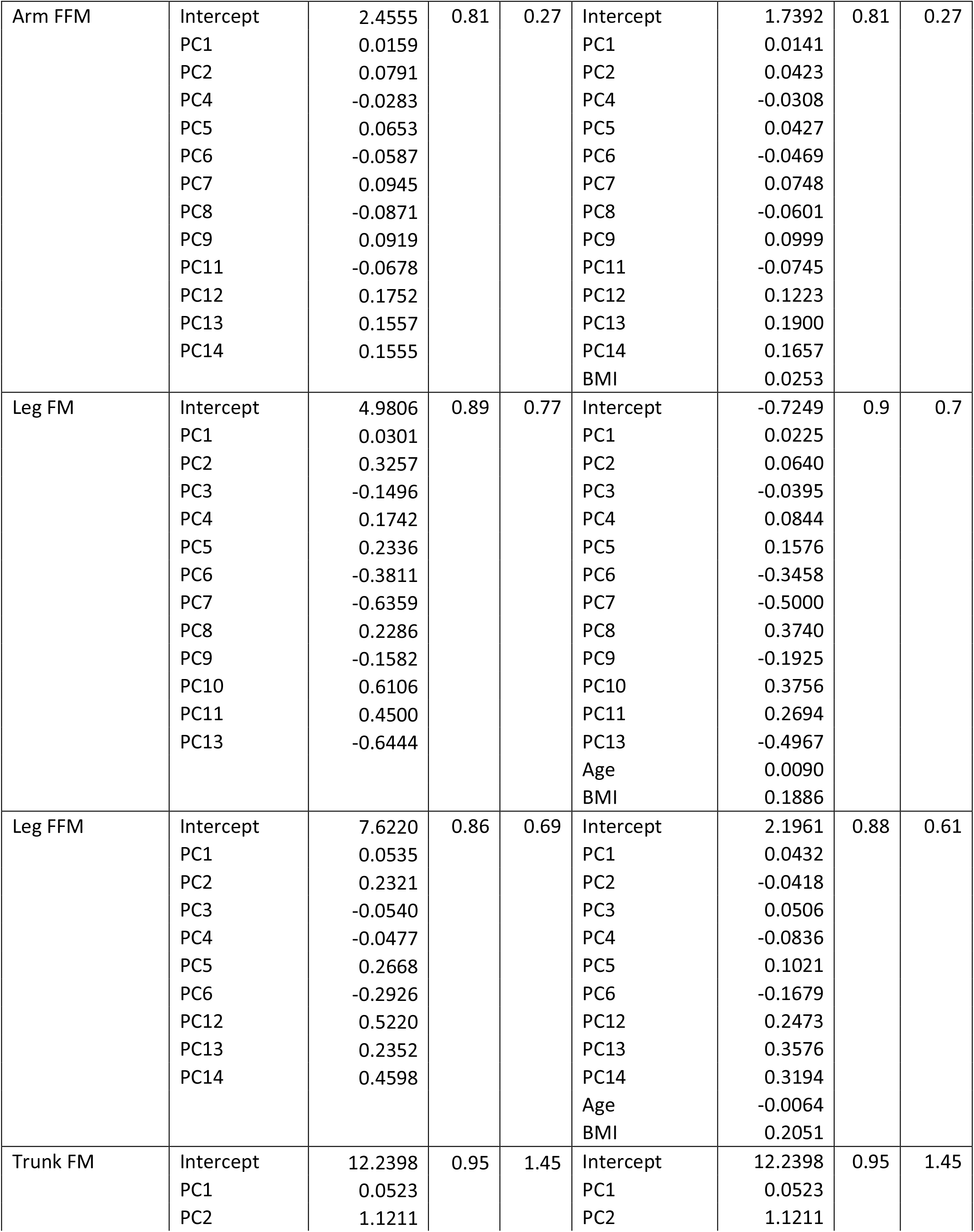
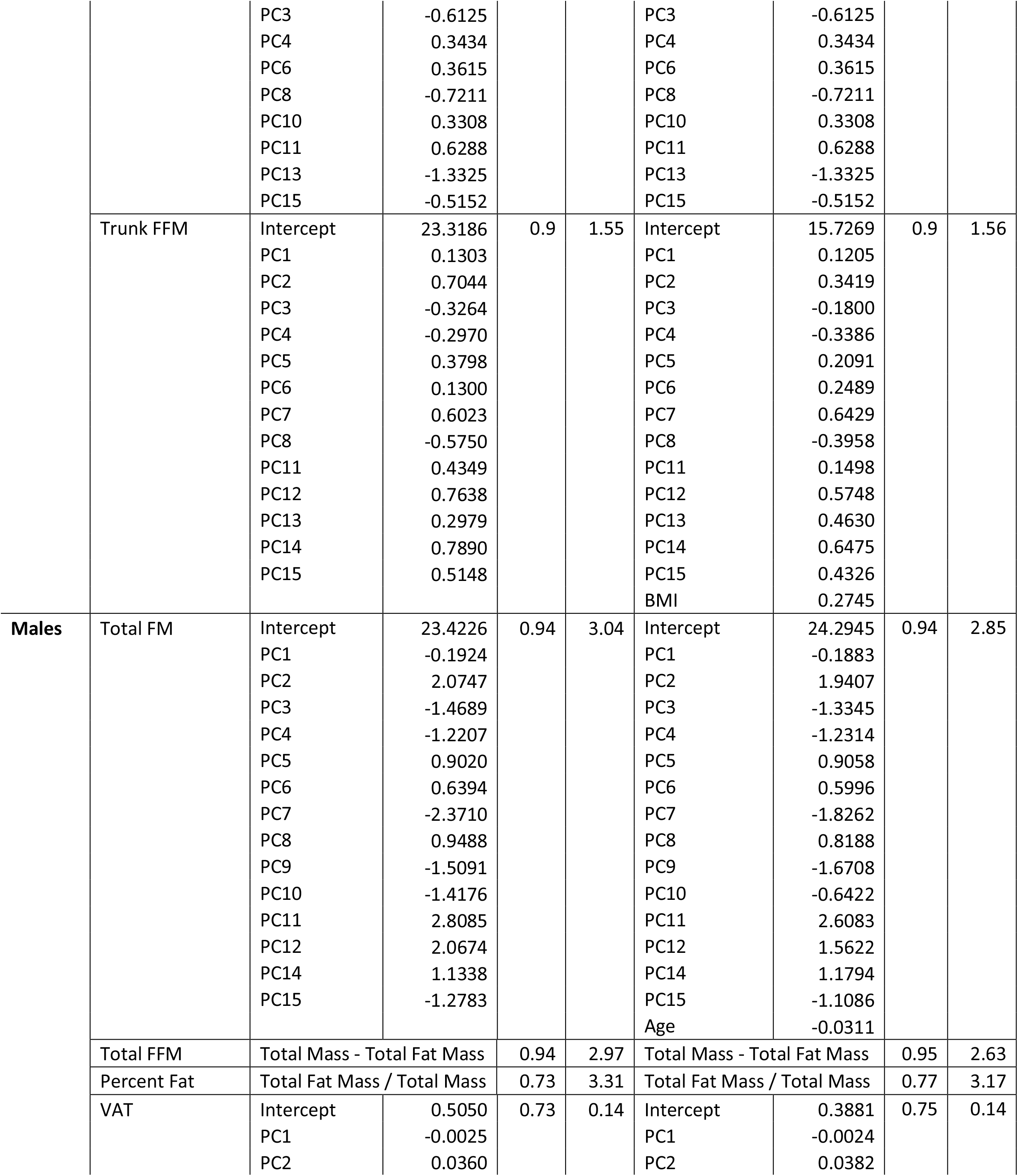
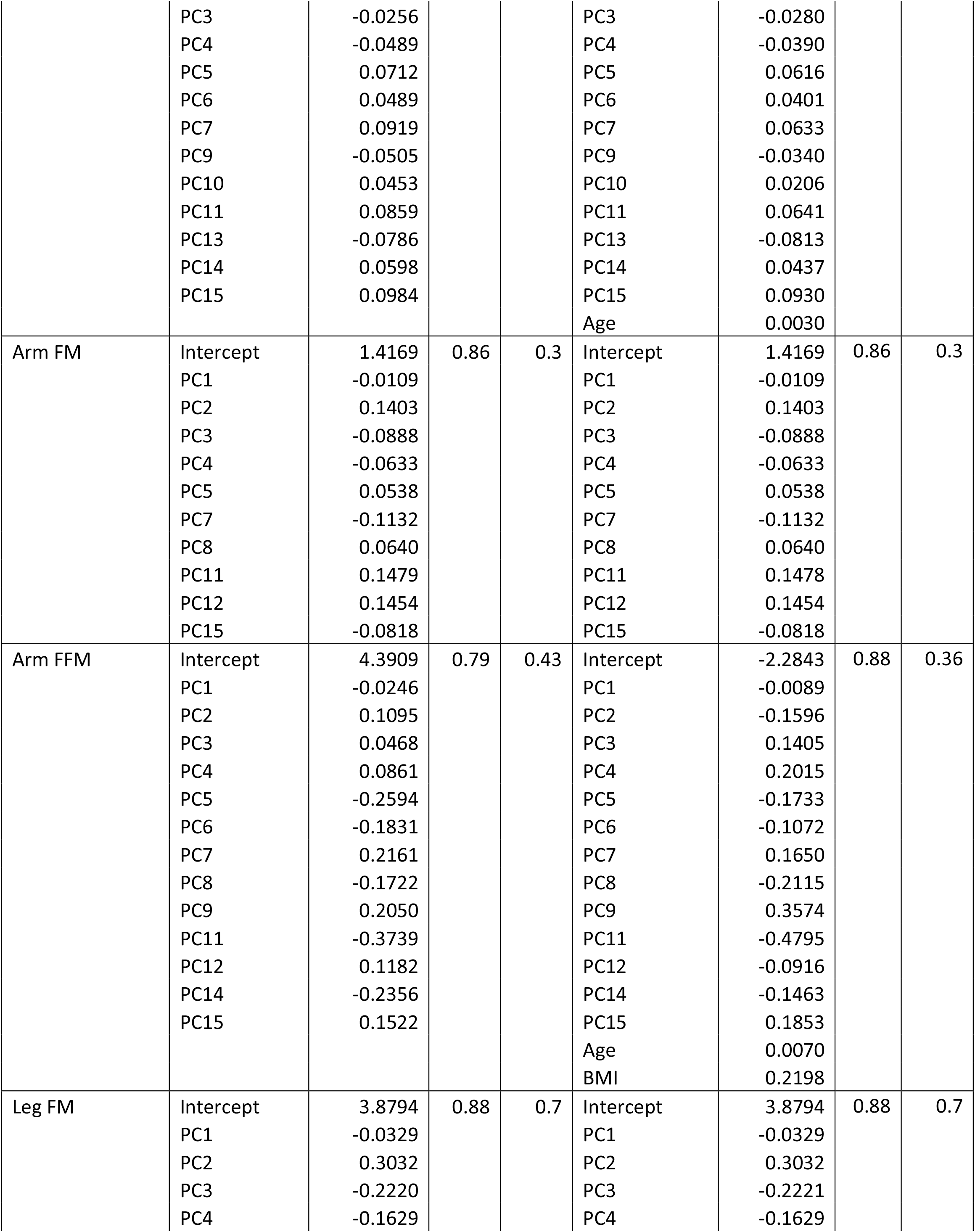
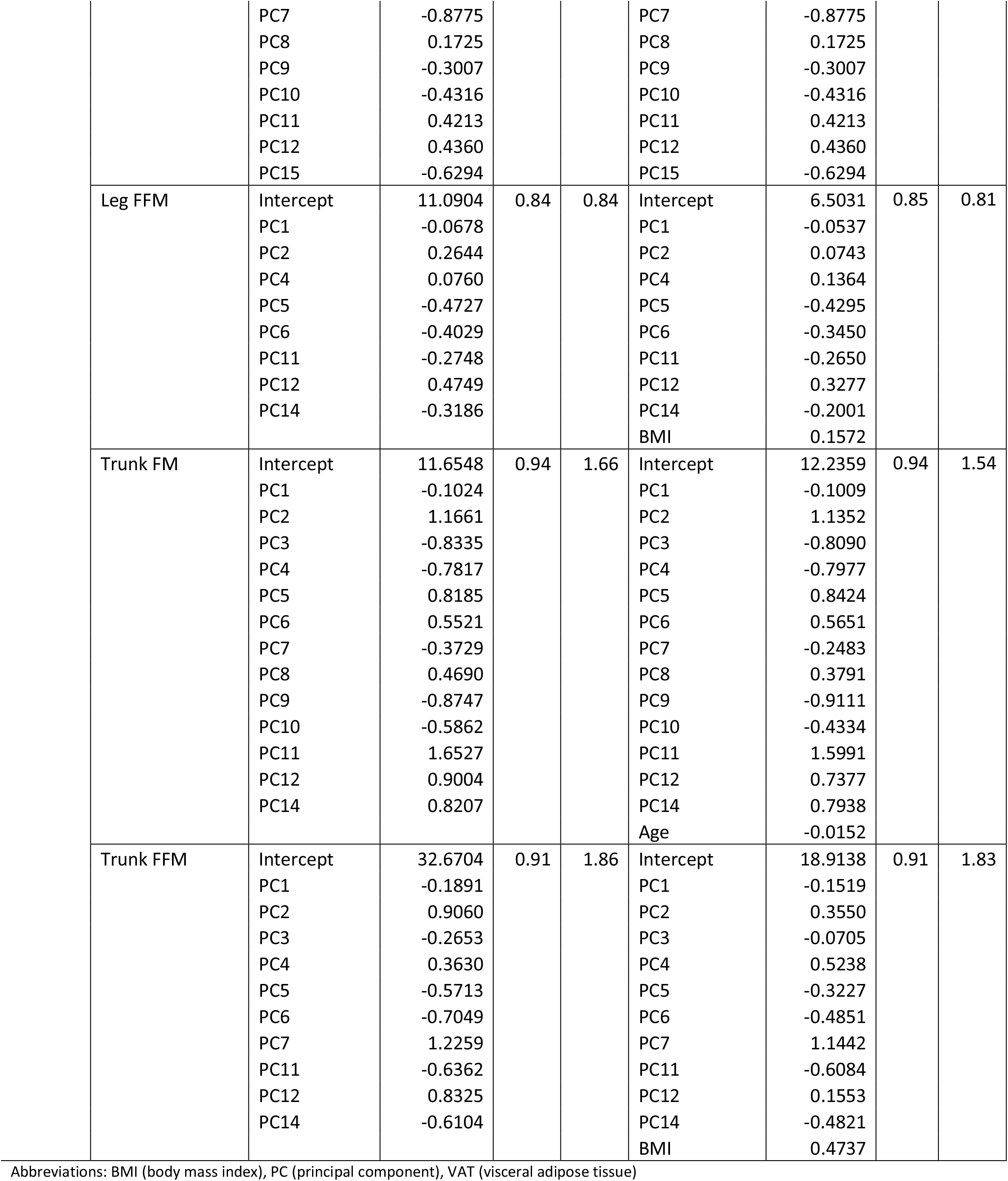
3DO body composition models to estimate to DXA body composition

|  |  | PC-only |  |  |  | PC + Demographics |  |  |  |
| --- | --- | --- | --- | --- | --- | --- | --- | --- | --- |
| Sex | Outcome | Variable | Coefficient | R<sup>2</sup> | RMSE | Variable | Coefficient | R<sup>2</sup> | RMSE |
| Females | Total FM | Intercept | 26.5689 | 0.94 | 2.91 | Intercept | 16.4494 | 0.94 | 2.88 |
|  |  | PC1 | 0.1282 |  |  | PC1 | 0.1108 |  |  |
|  |  | PC2 | 2.0930 |  |  | PC2 | 1.5903 |  |  |
|  |  | PC3 | -1.0575 |  |  | PC3 | -0.7970 |  |  |
|  |  | PC4 | 0.7487 |  |  | PC4 | 0.5483 |  |  |
|  |  | PC5 | 0.5329 |  |  | PC5 | 0.4035 |  |  |
|  |  | PC6 | -0.4188 |  |  | PC6 | -0.2978 |  |  |
|  |  | PC7 | -1.3200 |  |  | PC7 | -0.9192 |  |  |
|  |  | PC9 | -0.4824 |  |  | PC9 | -0.7256 |  |  |
|  |  | PC10 | 1.6580 |  |  | PC10 | 1.1085 |  |  |
|  |  | PC11 | 1.8068 |  |  | PC11 | 1.4661 |  |  |
|  |  | PC13 | -2.7090 |  |  | PC13 | -2.3351 |  |  |
|  |  |  |  |  |  | BMI | 0.3597 |  |  |
|  | Total FFM | Total Mass - Total Fat Mass |  | 0.92 | 2.76 | Total Mass - Total Fat Mass |  | 0.92 | 2.63 |
|  | Percent Fat | Total Fat Mass / Total Mass |  | 0.75 | 3.82 | Total Fat Mass / Total Mass |  | 0.75 | 3.76 |
|  | VAT | Intercept | 0.4516 | 0.72 | 0.15 | Intercept | 0.3334 | 0.78 | 0.14 |
|  |  | PC2 | 0.0408 |  |  | PC2 | 0.0431 |  |  |
|  |  | PC3 | -0.0226 |  |  | PC3 | -0.0254 |  |  |
|  |  | PC4 | 0.0151 |  |  | PC4 | 0.0142 |  |  |
|  |  | PC5 | -0.0332 |  |  | PC5 | -0.0252 |  |  |
|  |  | PC6 | 0.0554 |  |  | PC6 | 0.0417 |  |  |
|  |  | PC7 | 0.0349 |  |  | PC7 | 0.0382 |  |  |
|  |  | PC8 | -0.0680 |  |  | PC8 | -0.0931 |  |  |
|  |  | PC12 | -0.0809 |  |  | PC12 | -0.0301 |  |  |
|  |  | PC13 | -0.0348 |  |  | PC13 | -0.0569 |  |  |
|  |  | PC14 | -0.0489 |  |  | PC14 | -0.0555 |  |  |
|  |  | PC15 | -0.0570 |  |  | PC15 | -0.0490 |  |  |
|  |  |  |  |  |  | Age | 0.0028 |  |  |
|  | Arm FM | Intercept | 1.6889 | 0.86 | 0.34 | Intercept | 0.6964 | 0.87 | 0.33 |
|  |  | PC1 | 0.0073 |  |  | PC1 | 0.0058 |  |  |
|  |  | PC2 | 0.1508 |  |  | PC2 | 0.1023 |  |  |
|  |  | PC3 | -0.0723 |  |  | PC3 | -0.0508 |  |  |
|  |  | PC4 | 0.0354 |  |  | PC4 | 0.0242 |  |  |
|  |  | PC7 | -0.0391 |  |  | PC7 | -0.0176 |  |  |
|  |  | PC8 | -0.0928 |  |  | PC8 | -0.0755 |  |  |
|  |  | PC10 | 0.0857 |  |  | PC10 | 0.0553 |  |  |
|  |  | PC11 | 0.1248 |  |  | PC11 | 0.0901 |  |  |
|  |  | PC13 | -0.0635 |  |  | PC13 | -0.0259 |  |  |
|  |  | PC14 | 0.1282 |  |  | PC14 | 0.0937 |  |  |
|  |  |  |  |  |  | BMI | 0.0356 |  |  |

|  |  |  |  |  |  |  |  |  |
| --- | --- | --- | --- | --- | --- | --- | --- | --- |
| Arm FFM | Intercept | 2.4555 | 0.81 | 0.27 | Intercept | 1.7392 | 0.81 | 0.27 |
|  | PC1 | 0.0159 |  |  | PC1 | 0.0141 |  |  |
|  | PC2 | 0.0791 |  |  | PC2 | 0.0423 |  |  |
|  | PC4 | -0.0283 |  |  | PC4 | -0.0308 |  |  |
|  | PC5 | 0.0653 |  |  | PC5 | 0.0427 |  |  |
|  | PC6 | -0.0587 |  |  | PC6 | -0.0469 |  |  |
|  | PC7 | 0.0945 |  |  | PC7 | 0.0748 |  |  |
|  | PC8 | -0.0871 |  |  | PC8 | -0.0601 |  |  |
|  | PC9 | 0.0919 |  |  | PC9 | 0.0999 |  |  |
|  | PC11 | -0.0678 |  |  | PC11 | -0.0745 |  |  |
|  | PC12 | 0.1752 |  |  | PC12 | 0.1223 |  |  |
|  | PC13 | 0.1557 |  |  | PC13 | 0.1900 |  |  |
|  | PC14 | 0.1555 |  |  | PC14 | 0.1657 |  |  |
|  |  |  |  |  | BMI | 0.0253 |  |  |
| Leg FM | Intercept | 4.9806 | 0.89 | 0.77 | Intercept | -0.7249 | 0.9 | 0.7 |
|  | PC1 | 0.0301 |  |  | PC1 | 0.0225 |  |  |
|  | PC2 | 0.3257 |  |  | PC2 | 0.0640 |  |  |
|  | PC3 | -0.1496 |  |  | PC3 | -0.0395 |  |  |
|  | PC4 | 0.1742 |  |  | PC4 | 0.0844 |  |  |
|  | PC5 | 0.2336 |  |  | PC5 | 0.1576 |  |  |
|  | PC6 | -0.3811 |  |  | PC6 | -0.3458 |  |  |
|  | PC7 | -0.6359 |  |  | PC7 | -0.5000 |  |  |
|  | PC8 | 0.2286 |  |  | PC8 | 0.3740 |  |  |
|  | PC9 | -0.1582 |  |  | PC9 | -0.1925 |  |  |
|  | PC10 | 0.6106 |  |  | PC10 | 0.3756 |  |  |
|  | PC11 | 0.4500 |  |  | PC11 | 0.2694 |  |  |
|  | PC13 | -0.6444 |  |  | PC13 | -0.4967 |  |  |
|  |  |  |  |  | Age | 0.0090 |  |  |
|  |  |  |  |  | BMI | 0.1886 |  |  |
| Leg FFM | Intercept | 7.6220 | 0.86 | 0.69 | Intercept | 2.1961 | 0.88 | 0.61 |
|  | PC1 | 0.0535 |  |  | PC1 | 0.0432 |  |  |
|  | PC2 | 0.2321 |  |  | PC2 | -0.0418 |  |  |
|  | PC3 | -0.0540 |  |  | PC3 | 0.0506 |  |  |
|  | PC4 | -0.0477 |  |  | PC4 | -0.0836 |  |  |
|  | PC5 | 0.2668 |  |  | PC5 | 0.1021 |  |  |
|  | PC6 | -0.2926 |  |  | PC6 | -0.1679 |  |  |
|  | PC12 | 0.5220 |  |  | PC12 | 0.2473 |  |  |
|  | PC13 | 0.2352 |  |  | PC13 | 0.3576 |  |  |
|  | PC14 | 0.4598 |  |  | PC14 | 0.3194 |  |  |
|  |  |  |  |  | Age | -0.0064 |  |  |
|  |  |  |  |  | BMI | 0.2051 |  |  |
| Trunk FM | Intercept | 12.2398 | 0.95 | 1.45 | Intercept | 12.2398 | 0.95 | 1.45 |
|  | PC1 | 0.0523 |  |  | PC1 | 0.0523 |  |  |
|  | PC2 | 1.1211 |  |  | PC2 | 1.1211 |  |  |
|  |  | PC3 | -0.6125 |  |  | PC3 | -0.6125 |  |  |
|  |  | PC4 | 0.3434 |  |  | PC4 | 0.3434 |  |  |
|  |  | PC6 | 0.3615 |  |  | PC6 | 0.3615 |  |  |
|  |  | PC8 | -0.7211 |  |  | PC8 | -0.7211 |  |  |
|  |  | PC10 | 0.3308 |  |  | PC10 | 0.3308 |  |  |
|  |  | PC11 | 0.6288 |  |  | PC11 | 0.6288 |  |  |
|  |  | PC13 | -1.3325 |  |  | PC13 | -1.3325 |  |  |
|  |  | PC15 | -0.5152 |  |  | PC15 | -0.5152 |  |  |
|  | Trunk FFM | Intercept | 23.3186 | 0.9 | 1.55 | Intercept | 15.7269 | 0.9 | 1.56 |
|  |  | PC1 | 0.1303 |  |  | PC1 | 0.1205 |  |  |
|  |  | PC2 | 0.7044 |  |  | PC2 | 0.3419 |  |  |
|  |  | PC3 | -0.3264 |  |  | PC3 | -0.1800 |  |  |
|  |  | PC4 | -0.2970 |  |  | PC4 | -0.3386 |  |  |
|  |  | PC5 | 0.3798 |  |  | PC5 | 0.2091 |  |  |
|  |  | PC6 | 0.1300 |  |  | PC6 | 0.2489 |  |  |
|  |  | PC7 | 0.6023 |  |  | PC7 | 0.6429 |  |  |
|  |  | PC8 | -0.5750 |  |  | PC8 | -0.3958 |  |  |
|  |  | PC11 | 0.4349 |  |  | PC11 | 0.1498 |  |  |
|  |  | PC12 | 0.7638 |  |  | PC12 | 0.5748 |  |  |
|  |  | PC13 | 0.2979 |  |  | PC13 | 0.4630 |  |  |
|  |  | PC14 | 0.7890 |  |  | PC14 | 0.6475 |  |  |
|  |  | PC15 | 0.5148 |  |  | PC15 | 0.4326 |  |  |
|  |  |  |  |  |  | BMI | 0.2745 |  |  |
| Males | Total FM | Intercept | 23.4226 | 0.94 | 3.04 | Intercept | 24.2945 | 0.94 | 2.85 |
|  |  | PC1 | -0.1924 |  |  | PC1 | -0.1883 |  |  |
|  |  | PC2 | 2.0747 |  |  | PC2 | 1.9407 |  |  |
|  |  | PC3 | -1.4689 |  |  | PC3 | -1.3345 |  |  |
|  |  | PC4 | -1.2207 |  |  | PC4 | -1.2314 |  |  |
|  |  | PC5 | 0.9020 |  |  | PC5 | 0.9058 |  |  |
|  |  | PC6 | 0.6394 |  |  | PC6 | 0.5996 |  |  |
|  |  | PC7 | -2.3710 |  |  | PC7 | -1.8262 |  |  |
|  |  | PC8 | 0.9488 |  |  | PC8 | 0.8188 |  |  |
|  |  | PC9 | -1.5091 |  |  | PC9 | -1.6708 |  |  |
|  |  | PC10 | -1.4176 |  |  | PC10 | -0.6422 |  |  |
|  |  | PC11 | 2.8085 |  |  | PC11 | 2.6083 |  |  |
|  |  | PC12 | 2.0674 |  |  | PC12 | 1.5622 |  |  |
|  |  | PC14 | 1.1338 |  |  | PC14 | 1.1794 |  |  |
|  |  | PC15 | -1.2783 |  |  | PC15 | -1.1086 |  |  |
|  |  |  |  |  |  | Age | -0.0311 |  |  |
|  | Total FFM | Total Mass - Total Fat Mass |  | 0.94 | 2.97 | Total Mass - Total Fat Mass |  | 0.95 | 2.63 |
|  | Percent Fat | Total Fat Mass / Total Mass |  | 0.73 | 3.31 | Total Fat Mass / Total Mass |  | 0.77 | 3.17 |
|  | VAT | Intercept | 0.5050 | 0.73 | 0.14 | Intercept | 0.3881 | 0.75 | 0.14 |
|  |  | PC1 | -0.0025 |  |  | PC1 | -0.0024 |  |  |
|  |  | PC2 | 0.0360 |  |  | PC2 | 0.0382 |  |  |

|  |  |  |  |  |  |  |  |  |
| --- | --- | --- | --- | --- | --- | --- | --- | --- |
|  | PC3 | -0.0256 |  |  | PC3 | -0.0280 |  |  |
|  | PC4 | -0.0489 |  |  | PC4 | -0.0390 |  |  |
|  | PC5 | 0.0712 |  |  | PC5 | 0.0616 |  |  |
|  | PC6 | 0.0489 |  |  | PC6 | 0.0401 |  |  |
|  | PC7 | 0.0919 |  |  | PC7 | 0.0633 |  |  |
|  | PC9 | -0.0505 |  |  | PC9 | -0.0340 |  |  |
|  | PC10 | 0.0453 |  |  | PC10 | 0.0206 |  |  |
|  | PC11 | 0.0859 |  |  | PC11 | 0.0641 |  |  |
|  | PC13 | -0.0786 |  |  | PC13 | -0.0813 |  |  |
|  | PC14 | 0.0598 |  |  | PC14 | 0.0437 |  |  |
|  | PC15 | 0.0984 |  |  | PC15 | 0.0930 |  |  |
|  |  |  |  |  | Age | 0.0030 |  |  |
| Arm FM | Intercept | 1.4169 | 0.86 | 0.3 | Intercept | 1.4169 | 0.86 | 0.3 |
|  | PC1 | -0.0109 |  |  | PC1 | -0.0109 |  |  |
|  | PC2 | 0.1403 |  |  | PC2 | 0.1403 |  |  |
|  | PC3 | -0.0888 |  |  | PC3 | -0.0888 |  |  |
|  | PC4 | -0.0633 |  |  | PC4 | -0.0633 |  |  |
|  | PC5 | 0.0538 |  |  | PC5 | 0.0538 |  |  |
|  | PC7 | -0.1132 |  |  | PC7 | -0.1132 |  |  |
|  | PC8 | 0.0640 |  |  | PC8 | 0.0640 |  |  |
|  | PC11 | 0.1479 |  |  | PC11 | 0.1478 |  |  |
|  | PC12 | 0.1454 |  |  | PC12 | 0.1454 |  |  |
|  | PC15 | -0.0818 |  |  | PC15 | -0.0818 |  |  |
| Arm FFM | Intercept | 4.3909 | 0.79 | 0.43 | Intercept | -2.2843 | 0.88 | 0.36 |
|  | PC1 | -0.0246 |  |  | PC1 | -0.0089 |  |  |
|  | PC2 | 0.1095 |  |  | PC2 | -0.1596 |  |  |
|  | PC3 | 0.0468 |  |  | PC3 | 0.1405 |  |  |
|  | PC4 | 0.0861 |  |  | PC4 | 0.2015 |  |  |
|  | PC5 | -0.2594 |  |  | PC5 | -0.1733 |  |  |
|  | PC6 | -0.1831 |  |  | PC6 | -0.1072 |  |  |
|  | PC7 | 0.2161 |  |  | PC7 | 0.1650 |  |  |
|  | PC8 | -0.1722 |  |  | PC8 | -0.2115 |  |  |
|  | PC9 | 0.2050 |  |  | PC9 | 0.3574 |  |  |
|  | PC11 | -0.3739 |  |  | PC11 | -0.4795 |  |  |
|  | PC12 | 0.1182 |  |  | PC12 | -0.0916 |  |  |
|  | PC14 | -0.2356 |  |  | PC14 | -0.1463 |  |  |
|  | PC15 | 0.1522 |  |  | PC15 | 0.1853 |  |  |
|  |  |  |  |  | Age | 0.0070 |  |  |
|  |  |  |  |  | BMI | 0.2198 |  |  |
| Leg FM | Intercept | 3.8794 | 0.88 | 0.7 | Intercept | 3.8794 | 0.88 | 0.7 |
|  | PC1 | -0.0329 |  |  | PC1 | -0.0329 |  |  |
|  | PC2 | 0.3032 |  |  | PC2 | 0.3032 |  |  |
|  | PC3 | -0.2220 |  |  | PC3 | -0.2221 |  |  |
|  | PC4 | -0.1629 |  |  | PC4 | -0.1629 |  |  |
|  | PC7 | -0.8775 |  |  | PC7 | -0.8775 |  |  |
|  | PC8 | 0.1725 |  |  | PC8 | 0.1725 |  |  |
|  | PC9 | -0.3007 |  |  | PC9 | -0.3007 |  |  |
|  | PC10 | -0.4316 |  |  | PC10 | -0.4316 |  |  |
|  | PC11 | 0.4213 |  |  | PC11 | 0.4213 |  |  |
|  | PC12 | 0.4360 |  |  | PC12 | 0.4360 |  |  |
|  | PC15 | -0.6294 |  |  | PC15 | -0.6294 |  |  |
| Leg FFM | Intercept | 11.0904 | 0.84 | 0.84 | Intercept | 6.5031 | 0.85 | 0.81 |
|  | PC1 | -0.0678 |  |  | PC1 | -0.0537 |  |  |
|  | PC2 | 0.2644 |  |  | PC2 | 0.0743 |  |  |
|  | PC4 | 0.0760 |  |  | PC4 | 0.1364 |  |  |
|  | PC5 | -0.4727 |  |  | PC5 | -0.4295 |  |  |
|  | PC6 | -0.4029 |  |  | PC6 | -0.3450 |  |  |
|  | PC11 | -0.2748 |  |  | PC11 | -0.2650 |  |  |
|  | PC12 | 0.4749 |  |  | PC12 | 0.3277 |  |  |
|  | PC14 | -0.3186 |  |  | PC14 | -0.2001 |  |  |
|  |  |  |  |  | BMI | 0.1572 |  |  |
| Trunk FM | Intercept | 11.6548 | 0.94 | 1.66 | Intercept | 12.2359 | 0.94 | 1.54 |
|  | PC1 | -0.1024 |  |  | PC1 | -0.1009 |  |  |
|  | PC2 | 1.1661 |  |  | PC2 | 1.1352 |  |  |
|  | PC3 | -0.8335 |  |  | PC3 | -0.8090 |  |  |
|  | PC4 | -0.7817 |  |  | PC4 | -0.7977 |  |  |
|  | PC5 | 0.8185 |  |  | PC5 | 0.8424 |  |  |
|  | PC6 | 0.5521 |  |  | PC6 | 0.5651 |  |  |
|  | PC7 | -0.3729 |  |  | PC7 | -0.2483 |  |  |
|  | PC8 | 0.4690 |  |  | PC8 | 0.3791 |  |  |
|  | PC9 | -0.8747 |  |  | PC9 | -0.9111 |  |  |
|  | PC10 | -0.5862 |  |  | PC10 | -0.4334 |  |  |
|  | PC11 | 1.6527 |  |  | PC11 | 1.5991 |  |  |
|  | PC12 | 0.9004 |  |  | PC12 | 0.7377 |  |  |
|  | PC14 | 0.8207 |  |  | PC14 | 0.7938 |  |  |
|  |  |  |  |  | Age | -0.0152 |  |  |
| Trunk FFM | Intercept | 32.6704 | 0.91 | 1.86 | Intercept | 18.9138 | 0.91 | 1.83 |
|  | PC1 | -0.1891 |  |  | PC1 | -0.1519 |  |  |
|  | PC2 | 0.9060 |  |  | PC2 | 0.3550 |  |  |
|  | PC3 | -0.2653 |  |  | PC3 | -0.0705 |  |  |
|  | PC4 | 0.3630 |  |  | PC4 | 0.5238 |  |  |
|  | PC5 | -0.5713 |  |  | PC5 | -0.3227 |  |  |
|  | PC6 | -0.7049 |  |  | PC6 | -0.4851 |  |  |
|  | PC7 | 1.2259 |  |  | PC7 | 1.1442 |  |  |
|  | PC11 | -0.6362 |  |  | PC11 | -0.6084 |  |  |
|  | PC12 | 0.8325 |  |  | PC12 | 0.1553 |  |  |
|  | PC14 | -0.6104 |  |  | PC14 | -0.4821 |  |  |
|  |  |  |  |  | BMI | 0.4737 |  |  |

Abbreviations: BMI (body mass index), PC (principal component), VAT (visceral adipose tissue)

3DO test-retest precision was comparable to DXA (Table 3). 3DO total FM and FFM achieved a %CV (RMSE) of 1.76% (0.44 kg) and 1.97% (0.44 kg) in females, while DXA had a %CV (RMSE) of 0.98% (0.24 kg) and 0.59% (0.27 kg), respectively. In females, 3DO VAT, arm FM, and trunk FM (%CV; 4.4%, 2.72%, and 1.55%, RMSE; 0.02 kg, 0.04 kg, and 0.18 kg, respectively) achieved better test-retest precision than DXA (%CV; 8.08%, 3.16%, and 1.16%, RMSE; 0.03 kg, 0.05 kg, and 0.23 kg, respectively). Test-retest precision for males had similar statistics. However, only 3DO VAT was better than DXA in males. When comparing the precision across subgroups of age, BMI, and ethnicity/race, there were no significant differences (data not shown, p-value > 0.109).

**Table 3.** Test-retest precision of 3DO and DXA in %CV (RMSE).

| Variable | DXA Female | 3DO Female | DXA Male | 3DO Male |
| --- | --- | --- | --- | --- |
| Total FM, kg | 0.98 (0.24) | 1.76 (0.44) | 1.37 (0.28) | 2.93 (0.60) |
| Total FFM, kg | 0.59 (0.27) | 0.96 (0.44) | 0.55 (0.36) | 0.91 (0.60) |
| Percent Fat, % | NA (0.33) | NA (0.66) | NA (0.31) | NA (0.74) |
| VAT, kg | 8.08 (0.03) | 4.40 (0.02) | 6.88 (0.03) | 4.78 (0.02) |
| Arm FM, kg | 3.16 (0.05) | 2.72 (0.04) | 3.43 (0.04) | 3.99 (0.05) |
| Arm FFM, kg | 2.14 (0.05) | 2.57 (0.06) | 1.89 (0.08) | 1.94 (0.08) |
| Leg FM, kg | 1.47 (0.07) | 2.90 (0.14) | 2.34 (0.08) | 4.29 (0.14) |
| Leg FFM, kg | 1.13 (0.08) | 1.95 (0.14) | 1.18 (0.13) | 1.22 (0.13) |
| Trunk FM, kg | 2.04 (0.23) | 1.55 (0.18) | 2.42 (0.25) | 3.03 (0.31) |
| Trunk FFM, kg | 1.16 (0.26) | 1.52 (0.34) | 1.07 (0.34) | 1.20 (0.38) |

Abbreviations: %CV (percent coefficient of variation), FM (fat mass), FFM (fat-free mass), RMSE (root mean square error), VAT (visceral adipose tissue)

When comparing 3DO total FM, FFM, %fat, and VAT to DXA by age subgroups (Table 4), there were no mean differences in either females or males (p-value > 0.068). For the BMI subgroups (Table 5), underweight females had significant differences for total FM, FFM, and %fat (mean differences (MD) = 1.23 kg, -1.23 kg, and 3.12%, respectively, p-value < 0.014) as well as %fat for the females with obesity (MD = -0.79%, p-value = 0.023). There were no mean differences in the male BMI subgroups (p-value > 0.314). In the ethnicity/racial subgroups (Table 6), female Asians, blacks, and NHOPI subgroups had mean differences in total FM (MD = 0.64 kg, -0.81 kg, -1.55 kg%, respectively), FFM (MD = -1.6 kg, 1.7 kg, and 3 kg%, respectively), and %fat (MD = 1.38%, -1.11%, and -2.24%, respectively) (p-value < 0.017). Additionally, the male NHOPI subgroup had mean differences for total FM, FFM, and %fat (MD = -1.45 kg, 1.45 kg, and -1.88%, respectively, p-value < 0.038). If ethnicity/race was adjusted in the model, differences in ethnicity/race subgroups were no longer significant (p-value > 0.99). However, underweight females still had a mean difference (p-value = 0.029).

**Table 4.** 3D optical vs DXA body composition by age group.

| Sex | Groups | n | Variable | Mean Difference | % Mean Difference | P-Value |
| --- | --- | --- | --- | --- | --- | --- |
| Female | Young | 126 | Total FM | -0.28 | N.S. | 0.303 |
|  | Middle Aged | 119 | Total FM | 0.05 | N.S. | 0.828 |
|  | Senior | 89 | Total FM | -0.03 | N.S. | 0.901 |
|  | Young | 126 | Total FFM | 0.28 | N.S. | 0.303 |
|  | Middle Aged | 119 | Total FFM | -0.05 | N.S. | 0.828 |
|  | Senior | 89 | Total FFM | 0.03 | N.S. | 0.901 |
|  | Young | 126 | %Fat | -0.23 | N.S. | 0.559 |
|  | Middle Aged | 119 | %Fat | 0.19 | N.S. | 0.566 |
|  | Senior | 89 | %Fat | 0.04 | N.S. | 0.932 |
|  | Young | 126 | VAT | -0.01 | N.S. | 0.204 |
|  | Middle Aged | 119 | VAT | 0.02 | N.S. | 0.068 |
|  | Senior | 89 | VAT | -0.01 | N.S. | 0.438 |
| Male | Young | 127 | Total FM | -0.01 | N.S. | 0.960 |
|  | Middle Aged | 93 | Total FM | 0.25 | N.S. | 0.373 |
|  | Senior | 61 | Total FM | -0.36 | N.S. | 0.288 |
|  | Young | 127 | Total FFM | 0.01 | N.S. | 0.960 |
|  | Middle Aged | 93 | Total FFM | -0.25 | N.S. | 0.373 |
|  | Senior | 61 | Total FFM | 0.36 | N.S. | 0.288 |
|  | Young | 127 | %Fat | 0.03 | N.S. | 0.923 |
|  | Middle Aged | 93 | %Fat | 0.35 | N.S. | 0.261 |
|  | Senior | 61 | %Fat | -0.37 | N.S. | 0.384 |
|  | Young | 127 | VAT | -0.00 | N.S. | 0.893 |
|  | Middle Aged | 93 | VAT | 0.01 | N.S. | 0.508 |
|  | Senior | 61 | VAT | -0.01 | N.S. | 0.589 |

Abbreviations: FM (fat mass), FFM (fat-free mass), N.S. (not significant), %Fat (percent fat), VAT (visceral adipose tissue)
Mean differences = 3DO - DXA
FM, FFM, and VAT are measured in kg.

**Table 5.** 3D optical vs DXA body composition by BMI.

| Sex | Groups | n | Variable | Mean Difference | %Mean Difference | P-Value |
| --- | --- | --- | --- | --- | --- | --- |
| Female | Underweight | 24 | Total FM | **1.23** | **12** | **0.014** |
|  | Normal | 119 | Total FM | -0.01 | N.S. | 0.958 |
|  | Overweight | 94 | Total FM | -0.13 | N.S. | 0.610 |
|  | Obese | 96 | Total FM | -0.54 | N.S. | 0.099 |
|  | Underweight | 24 | Total FFM | **-1.23** | **-3.4** | **0.014** |
|  | Normal | 89 | Total FFM | 0.01 | N.S. | 0.958 |
|  | Overweight | 94 | Total FFM | 0.13 | N.S. | 0.610 |
|  | Obese | 96 | Total FFM | 0.54 | N.S. | 0.099 |
|  | Underweight | 24 | %Fat | **3.12** |  | **0.010** |
|  | Normal | 119 | %Fat | -0.10 | N.S. | 0.785 |
|  | Overweight | 94 | %Fat | -0.18 | N.S. | 0.616 |
|  | Obese | 96 | %Fat | **-0.79** |  | **0.023** |
|  | Underweight | 24 | VAT | -0.01 | N.S. | 0.772 |
|  | Normal | 119 | VAT | -0.01 | N.S. | 0.238 |
|  | Overweight | 94 | VAT | 0.01 | N.S. | 0.286 |
|  | Obese | 96 | VAT | 0.00 | N.S. | 0.988 |
| Male | Underweight | 3 | Total FM | 2.97 | N.S. | 0.155 |
|  | Normal | 80 | Total FM | 0.11 | N.S. | 0.701 |
|  | Overweight | 112 | Total FM | -0.06 | N.S. | 0.779 |
|  | Obese | 85 | Total FM | -0.16 | N.S. | 0.633 |
|  | Underweight | 3 | Total FFM | -2.97 | N.S. | 0.155 |
|  | Normal | 80 | Total FFM | -0.11 | N.S. | 0.701 |
|  | Overweight | 112 | Total FFM | 0.06 | N.S. | 0.779 |
|  | Obese | 85 | Total FFM | 0.16 | N.S. | 0.633 |
|  | Underweight | 3 | %Fat | 7.12 | N.S. | 0.168 |
|  | Normal | 80 | %Fat | 0.21 | N.S. | 0.601 |
|  | Overweight | 112 | %Fat | -0.06 | N.S. | 0.804 |
|  | Obese | 85 | %Fat | -0.29 | N.S. | 0.356 |
|  | Underweight | 3 | VAT | -0.04 | N.S. | 0.407 |
|  | Normal | 80 | VAT | -0.01 | N.S. | 0.347 |
|  | Overweight | 112 | VAT | 0.01 | N.S. | 0.380 |
|  | Obese | 85 | VAT | 0.00 | N.S. | 0.936 |

Abbreviations: FM (fat mass), FFM (fat-free mass), N.S. (not significant), %Fat (percent fat), VAT (visceral adipose tissue)
Mean differences = 3DO - DXA
FM, FFM, and VAT are measured in kg.

**Table 6.** 3D optical vs DXA body composition by ethnic/racial group.

| Sex | Groups | n | Variable | Mean Difference | %Mean Difference | P-Value |
| --- | --- | --- | --- | --- | --- | --- |
| Female | Asian | 80 | Total FM | **0.64** | **3.5** | **0.017** |
|  | Black | 70 | Total FM | **-0.81** | **-2.6** | **0.015** |
|  | Hispanic | 47 | Total FM | 0.36 | N.S. | 0.386 |
|  | NHOPI | 28 | Total FM | **-1.55** | **-5.4** | **0.003** |
|  | White | 107 | Total FM | 0.01 | N.S. | 0.980 |
|  | Asian | 80 | Total FFM | **-0.64** | **-1.6** | **0.017** |
|  | Black | 70 | Total FFM | **0.81** | **1.7** | **0.015** |
|  | Hispanic | 47 | Total FFM | -0.36 | N.S. | 0.386 |
|  | NHOPI | 28 | Total FFM | **1.55** | **3** | **0.003** |
|  | White | 107 | Total FFM | -0.01 | N.S. | 0.980 |
|  | Asian | 80 | %Fat | **1.38** |  | **0.006** |
|  | Black | 70 | %Fat | **-1.11** |  | **0.010** |
|  | Hispanic | 47 | %Fat | 0.63 | N.S. | 0.305 |
|  | NHOPI | 28 | %Fat | **-2.24** |  | **0.002** |
|  | White | 107 | %Fat | -0.01 | N.S. | 0.979 |
|  | Asian | 80 | VAT | -0.00 | N.S. | 0.908 |
|  | Black | 70 | VAT | -0.02 | N.S. | 0.189 |
|  | Hispanic | 47 | VAT | 0.01 | N.S. | 0.627 |
|  | NHOPI | 28 | VAT | -0.00 | N.S. | 0.942 |
|  | White | 107 | VAT | 0.01 | N.S. | 0.362 |
| Male | Asian | 71 | Total FM | 0.29 | N.S. | 0.360 |
|  | Black | 62 | Total FM | 0.42 | N.S. | 0.164 |
|  | Hispanic | 32 | Total FM | -0.65 | N.S. | 0.226 |
|  | NHOPI | 16 | Total FM | **-1.45** | **-6.5** | **0.038** |
|  | White | 98 | Total FM | -0.01 | N.S. | 0.958 |
|  | Asian | 71 | Total FFM | -0.29 | N.S. | 0.360 |
|  | Black | 62 | Total FFM | -0.42 | N.S. | 0.164 |
|  | Hispanic | 32 | Total FFM | 0.65 | N.S. | 0.226 |
|  | NHOPI | 16 | Total FFM | **1.45** | **1.9** | **0.038** |
|  | White | 98 | Total FFM | 0.01 | N.S. | 0.958 |
|  | Asian | 71 | %Fat | 0.50 | N.S. | 0.258 |
|  | Black | 62 | %Fat | 0.57 | N.S. | 0.093 |
|  | Hispanic | 32 | %Fat | -0.57 | N.S. | 0.374 |
|  | NHOPI | 16 | %Fat | **-1.88** |  | **0.013** |
|  | White | 98 | %Fat | -0.08 | N.S. | 0.789 |
|  | Asian | 71 | VAT | -0.01 | N.S. | 0.421 |
|  | Black | 62 | VAT | -0.03 | N.S. | 0.105 |
|  | Hispanic | 32 | VAT | 0.02 | N.S. | 0.159 |
|  | NHOPI | 16 | VAT | 0.02 | N.S. | 0.669 |
|  | White | 98 | VAT | 0.02 | N.S. | 0.159 |

Abbreviations: FM (fat mass), FFM (fat-free mass), N.S. (not significant), %Fat (percent fat), VAT (visceral adipose tissue)
Mean differences = 3DO - DXA
FM, FFM, and VAT are measured in kg.

3DO body composition values were associated with blood biomarkers and thigh strength similarly to DXA (Supplemental Table 1). Although some associations were considered statistically significant, the trend of the association were the same in all except for thigh strength in the underweight group.

## DISCUSSION

In this study, 3DO body composition accuracy and precision was evaluated at the subgroup level of age, BMI, and ethnicity/race. Accuracy of subgroups were compared to DXA, while precision was an intra-group comparison. The analysis showed mean differences among underweight females, NHOPI females and males, and Asian and black females, while all other groups had no mean differences. However, most of these differences were considered small or moderate. There were no significant differences for precision among the subgroups. The overall analysis supports the use of 3DO body composition in most subgroups. Those who fall under those subgroups with differences should be cautioned before use. Specific equations may be needed for those groups.

As a whole, the accuracy and precision were similar to previous work done by Wong et al. (26). Methods to develop the statistical shape model and the body composition equations were the exact same. Due to the power needed in each subgroup, splitting the total sample in this analysis into a representative training and test set by subgroup was not possible due to the low representations in some subgroups (i.e., underweight and NHOPI). However, the previous work showed how these methods were robust and validated strongly in its accuracy (26).

In the female underweight group, there was a large difference for total FM and moderate difference for FFM (%MD = 12% and -3.4%, respectively). Since the mean total FM in the underweight group was low (approximately 10 kg), the %MD was proportionately higher even with a small difference. Although the models were adjusted for BMI, they were not adjusted for ethnicity. Since female Asians, blacks, and NHOPI had mean differences as well, the ethnic differences could be driving some of the bias as 44% of the underweight group were either Asian or black. The remaining 56% of the underweight were either white or Hispanic, who did not observe mean differences. However, after adjustment for ethnicity/race, 3DO still overestimated total FM in underweight females, which means the model may not have seen enough underweight variance in the sample or other unique shape features from this group. The female subgroup with obesity also had a mean difference in %fat (−0.79%). However, this mean difference was very small especially in a group with the highest mean %fat (41%). Since the difference was small and the total FM and FFM were not significant, the %fat difference may have been due to chance. The largest differences were seen in the NHOPI group for both males and females. The differences may be driven by the low representation in comparison to other ethnic/racial groups. In addition, differences in shape can also be driving these differences. Certain shape characteristics in Asian, black, and NHOPI could be different from the majority of the group, which lead to under or overestimations.

In a previous paper that used descriptive shape characteristics by PCA, the authors showed that the PCs were correlated to metabolic blood serum markers (23). However, the prediction of the biomarkers with PCs had a low to moderate correlation. In the present analysis, 3DO predicted body composition values show similar correlations as DXA to metabolic blood serum markers (Supplemental Table 1). Although the strength of the association may differ, 3DO and DXA were within 5% for most of the correlations. In addition, the trend of the associations to blood serum biomarkers were the same. Even with mean differences between 3DO and DXA for certain subgroups (i.e., NHOPI and underweight), there can be confidence that 3DO body composition estimates have similar physiological associations as DXA and can be used in place of DXA if DXA is unavailable.

The strength of the study was the scrutiny at the subgroup level. Many body composition studies focused on the entire sample, have a homogenous sample, or were not powered to evaluate accuracy and precision with these many strata. This work builds upon the previous 3DO literature and was the first to evaluate accuracy and precision from subgroups. In addition, only a single equation was needed for the majority of the subgroups that were explored. A subgroup-specific equation may be needed for those with mean differences.

However, there were also limitations. Some subgroups were underrepresented (i.e., NHOPI and underweight). As a result, a representative split for a training and test set was not viable. The current analysis was completed with a healthy sample, so the results may not be applicable to patients with body composition altering diseases. Future work can focus on populations with diseases, younger populations (birth to 17 years), and more accessible 3DO methods such as apps from smartphones (35, 36).

## Conclusion

3DO body composition has come a long way in a short amount of time. This work further justifies the use of 3DO as a reliable method for body composition. Although some subgroups had mean differences from DXA, the majority of the subgroups were accurate. Beyond clinical settings, the accessibility and safety of these devices are appealing for those that want to monitor body composition frequently.

## Data Availability

All data produced in the present study are available upon reasonable request to the authors

https://shepherdresearchlab.org/request-data/

## Acknowledgements

We thank all the participants for graciously giving their time to be part of the studies, our collaborators at each site for their part in the study, Tyler Carter and Greg Moore at Fit3D for providing us the 3DO data, and Naureen Mahmood and Talha Zaman for providing us the application program interface to register and repose our 3DO data. The data underlying this study cannot be made publicly available because the data contains patient identifying information. Data is available from the Shape Up! Studies for researchers who meet the criteria for access to confidential data. For details and to request an application, please contact John Shepherd or visit www.shapeup.shepherdresearchlab.org.

## Author Contributions

MCW and JAS designed and conducted the research; MCW, BQ, and JAS were part of the data analysis; MCW, YEL, NNK, CM, AKG, GM, SBH, and JAS were in charge of their respective study recruitment and protocols; MCW and JAS drafted the manuscript and were responsible over the final content. All authors reviewed and approved the final manuscript.

